# Prevalence of HIV in patients hospitalized for COVID-19 and associated outcomes: a systematic review and meta-analysis

**DOI:** 10.1101/2020.07.03.20143628

**Authors:** Paddy Ssentongo, Emily S. Heilbrunn, Anna E. Ssentongo, Shailesh Advani, Vernon M. Chinchilli, Jonathan J. Nunez, Ping Du

## Abstract

**Objective:** To conduct a systematic review and meta-analysis of the prevalence of HIV in patients hospitalized for COVID-19 and delineating clinical outcomes including mortality.

**Design/Methods:** MEDLINE, SCOPUS, OVID, and Cochrane Library databases and medrxiv.org were searched from January 1st, 2020, to June 15th, 2020. Data were extracted from studies reporting the prevalence of HIV among hospitalized COVID-19 patients and their clinical outcomes.

Analyses were performed using random-effects models on log-transformed proportions and risk ratio estimates, and heterogeneity was quantified.

**Results:** A total of 144,795 hospitalized COVID-19 patients were identified from 14 studies in North America, Europe, and Asia. Median age was 55 years, and 66% were male. The pooled prevalence of HIV in COVID-19 patients was 1.22% [95% confidence interval (CI): 0.61%-2.43%)] translating to a 2-fold increase compared to the respective local-level pooled HIV prevalence in the general population of 0.65% (95% CI: 0.48%-0.89%). When stratified by country, the pooled HIV prevalence among COVID-19 patients in United States (1.43%, 95% CI: 0.98%–2.07%) was significantly higher compared to Spain (0.26%, 95% CI: 0.23%-0.29%) but was not different from China (0.99%, 95% CI: 0.25%-3.85%). The pooled mortality rate in HIV-positive patients hospitalized for COVID-19 was 14.1% (95% CI: 5.78%-30.50%) and was substantially higher in the United States compared to other countries.

**Conclusions:** The prevalence of HIV among COVID-19 patients appeared higher than the general population, suggesting a greater susceptibility to COVID-19 for PLWH. The pooled mortality rate is high, but the rates vary significantly across countries.

**Suggested Reviewers:** Nelson Sewankambo, MD, PhD

Makerere University College of Health Sciences

**Opposed Reviewers:** 

## Introduction

As of June 23^rd^, 2020, the number of confirmed cases of COVID-19 was approximately 9,237,640, with 474,609 deaths globally.^[1]^ Prior studies have identified that comorbidities such as cancer, cardiovascular disease, hypertension and heart failure all significantly increase the risk of infection and mortality from COVID-19.^[2, 3]^ In addition, there remains a concern of an increased risk of infection among those with compromised immune system, such as people living with human immunodeficiency virus (HIV) or AIDS (PLWHA). However, a large gap in the literature exists on the role of HIV in the susceptibility of infection with SARS-CoV-2 and severity of COVID-19.

There is a growing concern that the immunosuppressing nature of HIV [4] may make PLWH more susceptible to adverse COVID-related health outcomes. With approximately 37.9 million across the globe, it remains imperative to estimate the burden of COVID-19 and its clinical outcomes such as intensive care unit (ICU) admission, ventilation, and death among these individuals.^[4]^ The U.S. Department of Health and Human Services stated, “At the present time, we have no specific information about the risk of COVID-19 in people with HIV.”^[5]^ But, it is later mentioned that PLWH with low CD4 counts and those not on antiretroviral therapy have the greatest risk of getting severe forms of COVID-19. This meta-analysis was conducted to (1) determine the prevalence of HIV in hospitalized patients with COVID-19 and (2) determine if there is an increased risk of ICU admission, ventilation, and death in PLWH with COVID-19.

## Methods

### Information Source, Search Strategy and Study Selection

The present study has been registered with PROSPERO (registration ID: CRD42020187980). The present study is being reported in accordance with the reporting guidance provided in the Preferred Reporting Items for Systematic Reviews and Meta-Analyses (PRISMA) statement (see PRISMA checklist in Additional file 1).^[6]^ We searched PubMed, Scopus, OVID, Web of Science, Cochrane Library (from January 1^st^, 2020 to June 7^th^, 2020). We searched the grey or difficult to locate literature, including Google Scholar and Medrxiv. We performed hand-searching of the reference lists of included studies, relevant reviews, or other relevant documents. Studies reporting the prevalence of HIV infections in individuals hospitalized for COVID-19 were included. No limitations regarding study design, year of publication, country of publication, or language were identified. Articles published in other languages were appropriately translated and further included in the screening. The primary outcome was the prevalence of HIV. Secondary outcomes were mortality rates, ICU admissions, and mechanical ventilation in individuals with HIV and COVID-19. Predefined search terms determined by The Medical Subject Headings (MeSH) included multiple combinations of the following: “Human Immunodeficiency Virus” OR “HIV” AND “COVID-19” OR “Coronavirus.” The comprehensive list of studies found as a result of our initial search were transferred into Endnote, which further removed duplicate studies.

### Eligibility Criteria

Studies were selected according to the following criteria: participants, condition or outcome(s) of interest, study design and context.

1. ***Participants (population):*** We included studies involving patients hospitalized for COVID-19, regardless of age.
2. ***Condition or outcome(s) of interest:*** The primary outcome was the prevalence of HIV in hospitalized patients with COVID-19. Secondary outcomes were mortality rates, ICU admission rates, and mechanical ventilation rates of COVID-19 patients with HIV in comparison to COVID-19 patients without HIV.
3. ***Study design and context:*** Eligible studies will be randomized controlled trials, observational cohort (prospective or retrospective), cases series and case-control studies. We excluded case reports. Two reviewers (ESH and AES) initially screened titles and abstracts of all identified articles for eligibility. Criteria of inclusion included articles that reported the number of HIV-positive COVID-19 patients as well as the rates of ICU admission, mechanical ventilation, or death in HIV-positive COVID-19 patients.

### Data Extraction

After initially screening articles for inclusion based on titles and abstracts, two independent reviewers (ESH and AES) then screened full-text articles. Disagreements were resolved by discussion to meet a consensus, if necessary. In scenarios where consensus was not reached, a third reviewer (PS) was recruited in order to reach a consensus. We extracted the following information: year of publication, date of the study, sample size, number of HIV-positive COVID-19 patients, percentage of HIV-positive COVID-19 patients, the number of patients with HIV who did or did not die, the number of patients with HIV who did and did not require mechanical ventilation, the number of HIV-positive patients who did and did not require ICU admission. Relevant statistics, such as the relative risk, odds ratios or hazard ratios of mechanical ventilation, ICU admission, and death were extracted as well.

### Study quality assessment

Two reviewers (EH and AES) independently assessed the quality of the included studies. The Newcastle-Ottawa Scale (NOS) was utilized for the quality assessment of the included studies.^[7]^ NOS scale rates observational studies based on 3 parameters: selection, comparability between the exposed and unexposed groups, and exposure/outcome assessment. It assigns a maximum of 4 stars for selection, 2 stars for comparability, and 3 stars for exposure/outcome assessment. Studies with less than 5 stars were considered low quality, 5–7 stars moderate quality, and more than 7 stars high quality. In reporting results, we gave preference for results from clinical trials over observational studies.

### Data synthesis and statistical analysis

We adopted a narrative approach describing the number of studies, study settings, diagnostic criteria COVID-19, and, antiretroviral treatment, CD4 count, HIV or SARS-CoV-2 viral load, study-level patient demographics. The primary outcome was the pooled prevalence of HIV among patients hospitalized for COVID-19. We also computed the relative risk of ICU admissions, use of ventilation, and mortality associated with HIV in the cohort of patients hospitalized for COVID-19. The metaprop and metagen functions from the R package meta were used to calculate the pooled effect estimates using random-effects models.^[8]^ We applied the DerSimonian and Laird random-effects method to estimate the pooled between-study variance (heterogeneity).^[9]^ Individual and pooled estimates were graphically displayed using forest plots. A random-effects model assumes the observed estimates can vary across studies because of real differences in the effect in each study as well as sampling variability (chance). Between-study heterogeneity was assessed using *I*^2^ statistics, expressed as % (low (25%), moderate (50%), and high (75%) and Cochrane’s *Q* statistic (significance level < 0.05).^[10]^ To investigate the sources of heterogeneity, we conducted subgroup and meta-regression analyses using the country of the study, study-level mean or median age, gender, continent, and mean CD4 T cell counts as the regressors. We assessed potential ascertainment bias (as might be caused by publication bias) with funnel plots, by plotting the study effect size against standard errors of the effect size, and Egger’s test.^[11]^ All statistical analyses were performed with R software, version 3.4.3 (R, College Station, TX).

## Results

### Overview

As seen in **Figure 1**, we identified a total of 515 studies from six databases. 213 were determined to be duplicates, leaving 302 for the initial screening of titles and abstracts. 196 were then excluded based on the abstract. Of the 106 full-text articles screened, additional 91 excluded, leaving a total of 14 studies included for quantitative and qualitative analysis. These studies represented 5 countries: United States, China, Germany, Spain, and Italy. Diagnostic criteria for COVID-19 were clinical and laboratory confirmed by polymerase chain reaction (PCR). HIV diagnosis was retrieved from electronic medical records or past medical history. The articles that were ultimately included contained a total of 40,000 COVID-19 cases and the cumulative number of patients with HIV was 592. The median age of the patients included in the study was 55 years. Sixty-six percent were male (12 studies), the median CD4 count was 547 (5 studies), and 100% were currently taking antiretroviral therapy (4 studies) (**Table 1**),

**Table.**

| Author | Year of study | Study period | Study design | Country | City name | City of study population | # PLWHIV in City of study | HIV prevalence in city of study* | Sample size | Age (y) | % male | # HIV | % HIV | % HAART | CD4 Count Median | Study Quality Score |
| --- | --- | --- | --- | --- | --- | --- | --- | --- | --- | --- | --- | --- | --- | --- | --- | --- |
| Härter et al | 2020 | 3/11/20-4/17/20 | Case series | Germany | Ulm | 126,329 | - | - | 33 | 48 | 90 | 33 | 100 | 100 | 695 | 7 |
| Blanco et al | 2020 | - | Case series | Spain | Barcelona | 5,575,000 | - | - | 5 | 38 | 60 | 5 | 100 | 100 | 564 | 7 |
| Gervason et al | 2020 | 2/21/20-4/20/20 | Cohort | Italy | Milano | 1,352,000 | - | - | 47 | 51 | 77 | 47 | 100 |  | 636 | 7 |
| Richardson et al | 2020 | 3/1/20-4/4/20 | Case series | United States | New York City | 8,000,000 | 108,000 | 1.35 | 5,700 | 63 | 60 | 43 | 0.75 | N/A | N/A | 8 |
| Chen et al | 2020 | 1/1/20-2/10/20 | Cohort | China | Wuhan | 9,000,000 | 6,000 | 0.067 | 203 | 54 | 53 | 2 | 0.99 | N/A | N/A | 8 |
| Karmen-tuohy et al | 2020 | 3/2/20-4/23/20 | Cohort | United States | New York City | 8,000,000 | 108,000 | 1.35 | 63 | - | - | 21 | 33.33 | N/A | N/A | 8 |
| Bhatraju et al | 2020 | Through 3/23/20 | Case series | United States | Seattle | 4,000,000 | 8,616 | 0.215 | 24 | 64 | 63 | 1 | 4.17 | N/A | N/A | 8 |
| Cummings et al | 2020 | 3/2/20-4/1/20 | Cohort | United States | New York City | 8,000,000 | 108,000 | 1.35 | 257 | 62 | 67 | 8 | 3.11 | N/A | N/A | 8 |
| Prieto-alhambra et al | 2020 | 3/15/20-4/24/20 | Cohort | Spain | Catalonia | 7,000,000 | 34,729 | 0.496 | 121,263 | - | 42 | 311 | 0.26 | N/A | N/A | 8 |
| Argenziano et al | 2020 | 3/1/20-4/5/20 | Cohort | United States | New York City | 8,000,000 | 10,8000 | 1.35 | 1,000 | 63 | 60 | 21 | 2.1 | N/A | N/A | 8 |
| Marcello et al | 2020 | 3/5/20-4/9/20 | Cohort | United States | New York City | 8,000,000 | 108,000 | 1.35 | 13,442 | 52.7 | 56 | 159 | 1.18 | N/A | N/A | 8 |
| Crotty et al | 2020 | 3/1/20-4/30/20 | Cohort | United States | Dallas | 2,500,000 | 16,825 | 0.673 | 289 | 58.6 | - | 7 | 2.42 | N/A | N/A | 8 |
| Suwanwongs e & shabarek et al | 2020 |  | Case Series | United States | New York City | 8,000,000 | 108,000 | 1.35 | 9 | 57.2 | 78 | 9 | 100 | 100 | 617 | 7 |
| Vizcarra et al | 2020 | Through 4/30/20 | Cohort | Spain | Madrid | 6,642,000 | - | - | 51 | 53.3 | 84 | 51 | 100 | 100 | 224 | 7 |

\* The city-level prevalence of HIV was calculated for the COVID-19 studies that we could calculate prevalence of HIV among COVID-19 patients
Abbreviations: PLWHIV: people living with HIV; HAART: Highly active antiretroviral therapy

**Figure 1:**
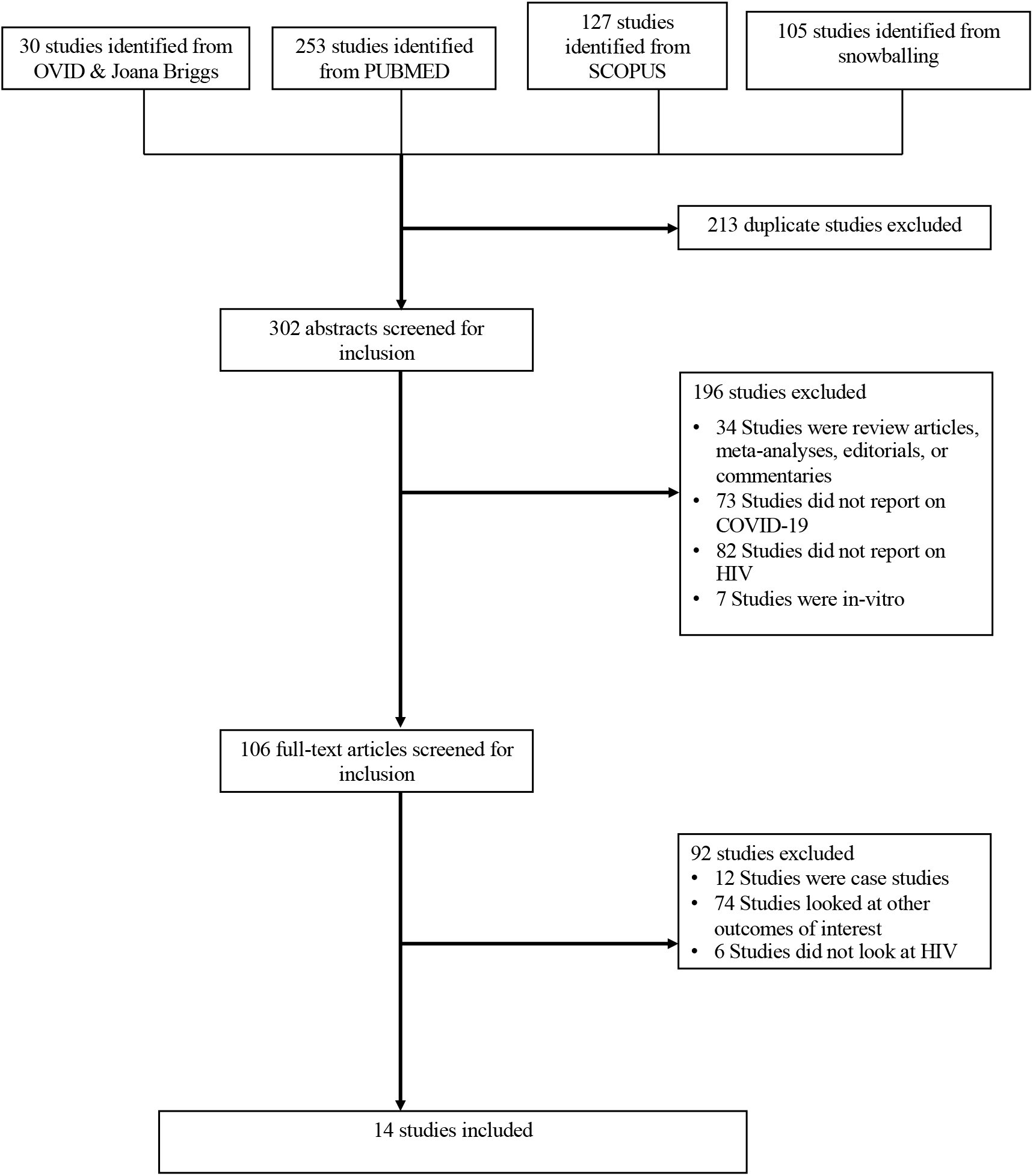
PRISMA flow diagram of the included articles in meta-analysis.

### Pooled prevalence of HIV among COVID-19 patients

The pooled prevalence of HIV in COVID-19 patients was 1.22 % [95% confidence interval (CI): 0.61%-2.43%)] (**Figure 2a**). Between-study heterogeneity for the prevalence was high (I^2^=98%; p<0·01). The prevalence of HIV in COVID-19 patients ranged from a low of 0.26 % (95% CI: 0.23%-0.29%) in Catalonia, Spain to a high of 4.17% (95% CI: 0.58%-24.35%) in Seattle, USA. We compared the prevalence of city-specific prevalence of HIV in COVID-19 with the corresponding population-level prevalence of HIV. The point estimates for the prevalence of HIV of the general population in the analyzed cities was half the HIV prevalence in among COVID-19 patients: 0.65% (95% CI: 0.48%-0.89%, **Figure 2b**), although not significantly different. When we stratified the analysis by the country (**Figure 3**), we found a noticeable difference in the pooled HIV prevalence among COVID-19 patients in United Sates (1.43%, 95% CI: 0.98%-2.07%) compared to Spain (0.26%, 95% CI: 0.23%-0.29%) but the difference was not significant compared to the prevalence in China (0.99%, 95% CI: 0.25 %-3.85%).

**Figure 2:**
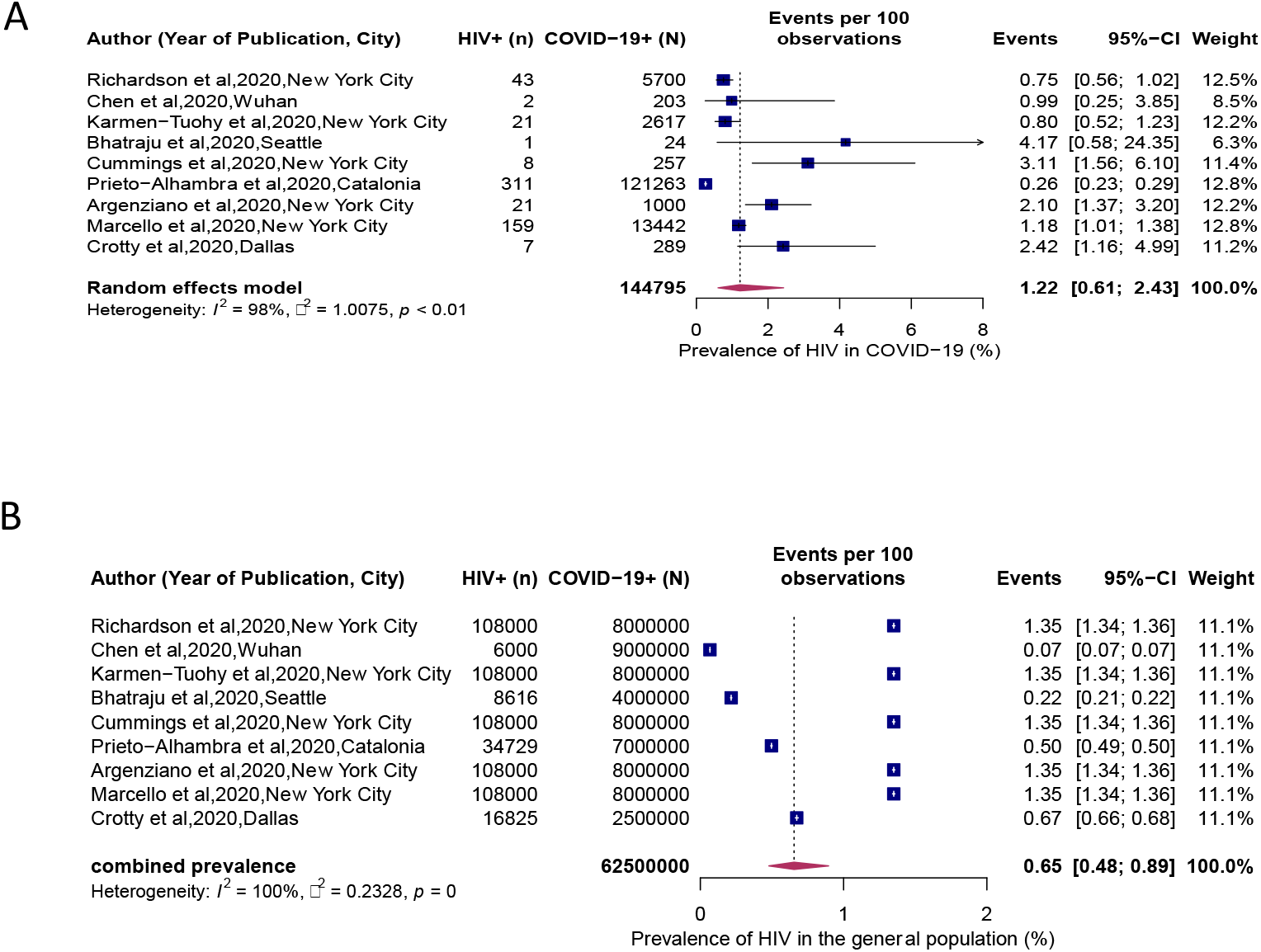
**(A) Prevalence of HIV in patients hospitalized for COVID-19. (B) Local Prevalence of HIV in cities where COVID-19 studies were conducted**

**Figure 3:**
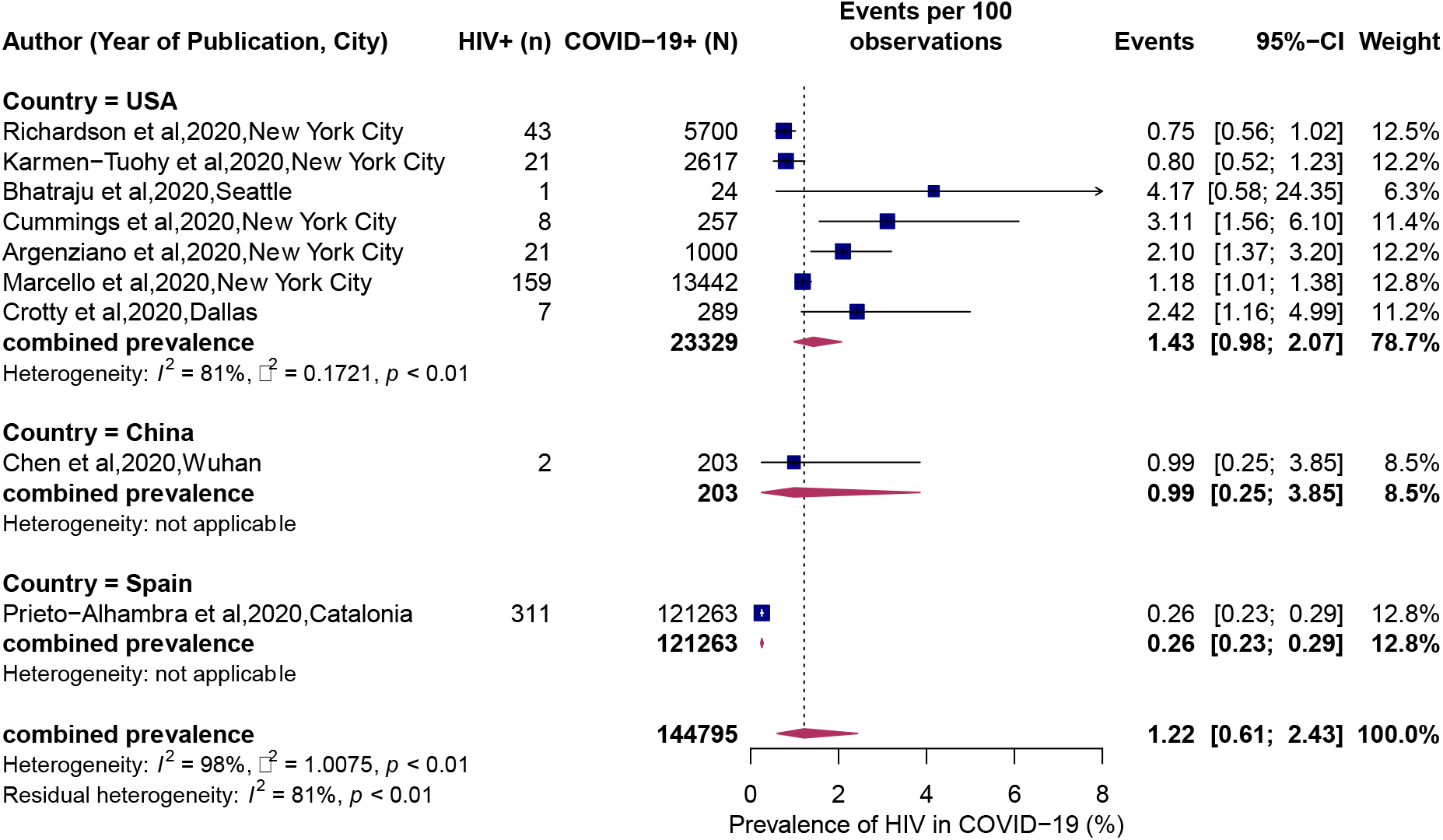
**Prevalence of HIV in patients hospitalized for COVID-19 stratified by country**

### Association of HIV and COVID-19 Clinical Outcomes

Five studies reported the mortality rates among individuals with HIV hospitalized for COVID-

19. The pooled mortality rate among HIV positive patients was 12.35% (95% CI 6.77%-21.49%, **Figure 4a)**. Between-study heterogeneity for the prevalence was low (I^2^=47%; p=11). The pooled prevalence ranged from a high of 35% (95% CI: 9.81%-72%) in the USA to a low of 3% (95% CI: 0.3%-20%, **Figure 4b**). Only two studies reported the strength of the association in the mortality rates between COVID-19 with HIV and COVID-19 patients without HIV. Therefore, it was most appropriate to report these values separately. The risk ratio of death in COVID-19 patients with HIV in a study conducted by Marcello and colleagues was 0.38 (95% CI 0.26, 0.55). On the contrary, the risk ratio of death in COVID-19 patients with HIV in a study conducted by Karmen-Tuohy and colleagues was 1.28 (95% CI 0.57, 2.90, **Supplemental Figure 1**). Similarly, the only two studies that reported on the association of ICU admission with COVID-19, found a non-significant higher risk (**Supplemental Figure 2**). The one study which explored the rates of mechanical ventilation between HIV positive and negative COVID-19, also found a non-significant higher risk in the HIV group. (RR: 2.31 (95% CI: 0.59-9.11)

**Figure 4:**
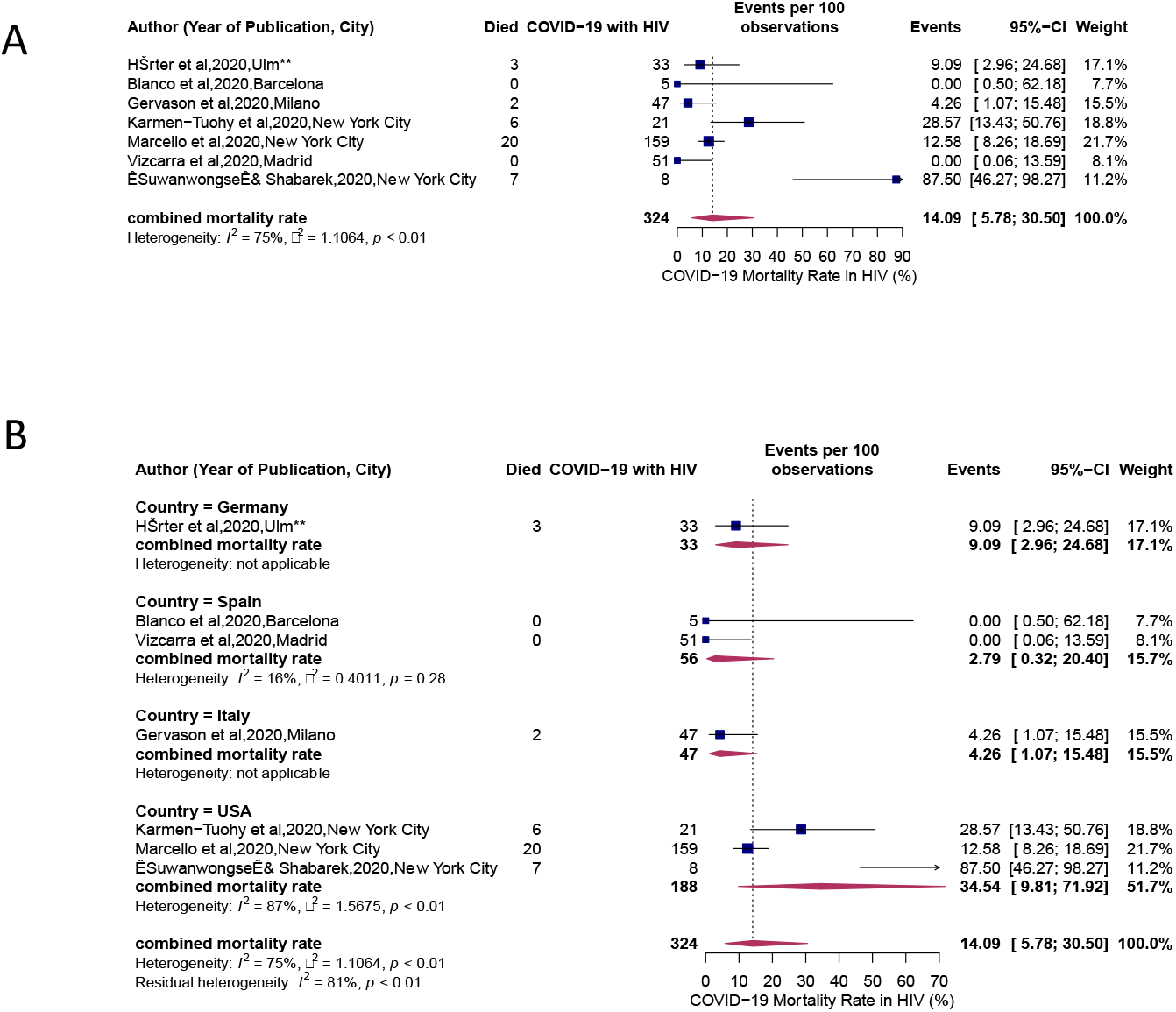
**(A) Mortality rates in HIV patients hospitalized COVID-19. (B) Mortality rates in HIV patients hospitalized COVID-19 stratified by country**

### Study quality, publication bias and sensitivity analyses

The median study quality score was 8 out of 9 (range=7-8, **Table 1**). Visual inspection of funnel plot shows clear evidence of heterogeneity and funnel plot asymmetry (**Supplementa**l **Figure 3**). Egger’s test for asymmetry was significant (p = 0.0052).

## Discussion

To our knowledge, our study remains one of the first to estimate the burden of COVID-19 among PLWH by conducting the meta-analysis of available studies. We found that the pooled prevalence of HIV among hospitalized patients with COVID-19 was 1.22% (95% CI=0.61, 2.43), while the mortality rate among hospitalized COVID-19 patients with HIV was 12.35% (95% CI=6.77, 21.49%). As this pandemic spreads globally and we continue to expand our understanding of the impact of COVID-19 has on patients with pre-existing conditions, including HIV, we found that estimates of both prevalence and mortality among HIV patients vary across these regions. Given the highly infectious nature of COVID-19 and its association with preexisting conditions, PLWH, even on antiretroviral therapy, may still present with a compromised immune system, and subsequently, have an increased risk of having COVID-19 and associated adverse outcomes.

In addition to immunosuppression that increases the risk of developing AIDS or non-AIDS-related infections, PLWH are at risk of anemia, neutropenia, thrombocytopenia, and abnormal serum electrolytes, which also plan an important role in COVID-19 disease course. Hence, it remains vital for PLWH to retain in regular HIV care and maintain good adherence to antiretroviral therapy. With this being said, PLWH on effective ART treatments also experience numerous non-AIDS noncommunicable diseases, such as hypertension and cardiovascular diseases). These comorbidities also play a significantly influential role in COVID-19 infection [2, 3]. Preliminary evidence from cohort studies suggest that patients with COVID-19 present with a dysregulated immune system characterized by high neutrophil to lymphocyte (NLR) ratio and CD4 T-cell lymphopenia. ^[12]^ The mean of age of HIV patients with COVID-19 was 55 years, which is a decade younger in comparison to the mean age of hospitalized COVID-19 patients in the general population. This large difference in chronological age could be due to the effect of premature aging of PLWH due to chronic inflammation that is resulted from HIV infection or high prevalence of certain behavioral risk factors (e.g., smoking). As a result of this, the rate of comorbidities that are commonly seen in older, non-HIV populations (such as cardiovascular disease) can happen in PLWH at a much younger age.^[13]^

PLWH are generally characterized by a dysregulated immune system. A rapid and well-coordinated innate immune response provides a vital defense system against viral infections. ^[14]^ It remains crucial to maintain a balance in the immune system, characterized by a balance in CD4 cell subpopulations that consist of effector-T cells and memory T-cells. Any imbalance can result in the weakening of the immune system, characterized by activation, proliferation, and effector functions of number cells leading to disturbance on homeostasis ^[12, 15]^. Among patients with COVID-19, evidence exists of high levels of proinflammatory cytokines and chemokines leading to increase in severity of COVID-19 infections, higher consumption of CD4+ and CD8+ T-cells, decrease in regulatory T-cells, and a disturbed innate immune environment leading to a cytokine storm and worsen damaged tissue ^[12, 16]^. In addition, evidence also suggests that subgroups of COVID-19 patients might suffer from hyperinflammatory syndrome termed as secondary hemophagocytic lymphohistiocytosis (sHLH) which is often triggered by underlying viral infections or sepsis and leads in fulminant and fatal hypercytokinaemia with multiorgan failure. ^[16]^ Though incidence of sHLH among patients with HIV might be rare, co-infection of HIV patients with COVID-19 might result in sHLH leading to heightened severity of disease and high mortality.

An important component and determinant of HIV infections involves not only CD4 count, but also viral load, and access and adherence to antiretroviral therapy which remains an important aspect of long-term outcomes, including progression to AIDS and survival among HIV patients.^[17]^ Non-adherence to ART have shown to be associated with low CD4 count, high viral load, and risk of AIDS and death. ^[18]^ Though our study could not identify the prevalence of COVID-19 infection stratified by their CD4 count or treatment regimens, these factors might have an indirect impact on COVID-19 infections and associated outcomes among PLWH.

Globally, HIV/AIDS has led to approximately 39 million deaths to date and 36 million people are living with HIV currently, with more than 2 million new infection diagnosed annually. ^[19]^ In contrast to patterns observed for COVID-19, there exists a higher prevalence and mortality of HIV among regions of Sub-Saharan Africa and Asia, among younger individuals in ages 15-49, among men and those countries with poor ART coverage. ^[20]^ The global burden of COVID-19 is rapidly evolving. Currently the United States, Brazil, United Kingdom, and India have highest prevalence of COVID-19 cases in the world. In terms of mortality, highest rates of death associated with COVID-19 have been observed in the United States, Brazil, UK, Italy, France, and Spain. In addition, COVID-19 infections have been seen among individuals with older age or those with comorbid conditions. Hence, future studies should further characterize the true burden of COVID-19 among PLWH by age, gender, socioeconomic status, geographic location, CD4 count, ART adherence, viral load, and comorbidities.

Our study has several strengths and limitations. We provide one of the first evidence of prevalence and mortality among hospitalized COVID-19 patients. Further, we incorporated all available literature to synthesize estimates of the burden of COVID-19 across different regions. However, the contrasting nature and limited evidence of COVID-19 infection rates by comorbidities across regions from Asia and Africa may limit the generalizability of our true estimates. Further, as many studies do not report on the HIV disease status based on their CD4 count and treatment regimen, we could not estimate the severity of COVID-19 infection among HIV positive individuals.

## Conclusion

In conclusion, the prevalence of HIV among hospitalized COVID-19 patients appeared to be higher compared to the general population, suggesting a greater susceptibility to COVID-19. The mortality rates remain high which highlights the need to develop targeted intervention programs, including medications, mental health services, health education, use of telemedicine, timely isolation, and contact tracing among individuals with immunocompromised states to reduce the health impact of COVID-19 pandemic on those individuals. In addition, among regions with high burden of HIV, further studies are warranted to estimate the impact of COVID-19 by increasing testing capacity and assessing long term outcomes.

## Data Availability

All data is found within the manuscript.

## Ethics approval

Not applicable

## Consent for publication

No consent to publish was needed for this study as we did not use any details, images or videos related to individual participants.

## Authors’ contributions

PS has full access to all of the data in the study and takes responsibility for the integrity of the data and the accuracy of the data analysis.

Concept and design: PS, PD

Data acquisition, analysis and interpretation: PS, AES, ESH, SA, VMC JJN, PD Writing the manuscript: PS, ESH, SA

Statistical analysis: PS, VMC,

PD Supervision: PD

All authors read and approved the final version of this manuscript.

## Acknowledgment

None

**Supplemental Data File (.doc, .tif, pdf, etc.)**

Supplemental documents.doc

## References

1. Johns Hopkins University. CORONAVIRUS RESOURCE CENTER. In; 2020.

2. Ssentongo P, Ssentongo AE, Heilbrunn ES, Chinchilli VM. The association of cardiovascular disease and other pre-existing comorbidities with COVID-19 mortality: A systematic review and meta-analysis. medRxiv 2020.

3. Guan W-j, Liang W-h, Zhao Y, Liang H-r, Chen Z-s, Li Y-m, et al. Comorbidity and its impact on 1590 patients with Covid-19 in China: A Nationwide Analysis. European Respiratory Journal 2020; 55(5).

4. Frank TD, Carter A, Jahagirdar D, Biehl MH, Douwes-Schultz D, Larson SL, et al. Global, regional, and national incidence, prevalence, and mortality of HIV, 1980–2017, and forecasts to 2030, for 195 countries and territories: a systematic analysis for the Global Burden of Diseases, Injuries, and Risk Factors Study 2017. The Lancet HIV 2019.

5. HIV.GOV. Coronavirus (COVID-19) and People with HIV. In; 2020.

6. Moher D, Liberati A, Tetzlaff J, Altman DG, Group P. Preferred reporting items for systematic reviews and meta-analyses: the PRISMA statement. PLoS med 2009; 6(7):e1000097.

7. Peterson J, Welch V, Losos M, Tugwell P. The Newcastle-Ottawa scale (NOS) for assessing the quality of nonrandomised studies in meta-analyses. Ottawa: Ottawa Hospital Research Institute 2011.

8. Schwarzer G, Carpenter JR, Rücker G. Meta-analysis with R. Springer; 2015.

9. DerSimonian R, Kacker R. Random-effects model for meta-analysis of clinical trials: an update. Contemporary clinical trials 2007; 28(2):105–114.

10. Higgins JP, Thompson SG, Deeks JJ, Altman DG. Measuring inconsistency in meta-analyses. Bmj 2003; 327(7414):557–560.

11. Egger M, Smith GD, Schneider M, Minder C. Bias in meta-analysis detected by a simple, graphical test. Bmj 1997; 315(7109):629–634.

12. Qin C, Zhou L, Hu Z, Zhang S, Yang S, Tao Y, et al. Dysregulation of immune response in patients with COVID-19 in Wuhan, China. Clinical Infectious Diseases 2020.

13. Maciel RA, Klück HM, Durand M, Sprinz E. Comorbidity is more common and occurs earlier in persons living with HIV than in HIV-uninfected matched controls, aged 50 years and older: a cross-sectional study. International Journal of Infectious Diseases 2018; 70:30–35.

14. Shaw AC, Goldstein DR, Montgomery RR. Age-dependent dysregulation of innate immunity. Nature Reviews Immunology 2013; 13(12):875–887.

15. Moro-García MA, Alonso-Arias R, López-Larrea C. When aging reaches CD4+ T-cells: phenotypic and functional changes. Frontiers in immunology 2013; 4:107.

16. Mehta P, McAuley DF, Brown M, Sanchez E, Tattersall RS, Manson JJ, et al. COVID-19: consider cytokine storm syndromes and immunosuppression. Lancet (London, England) 2020; 395(10229):1033.

17. Kaplan JE, Hanson DL, Jones JL, Dworkin MS. Viral load as an independent risk factor for opportunistic infections in HIV-infected adults and adolescents. Aids 2001; 15(14):1831–1836.

18. Sherr L, Lampe F, Clucas C, Johnson M, Fisher M, Leake Date H, et al. Self-reported non-adherence to ART and virological outcome in a multiclinic UK study. AIDS care 2010; 22(8):939–945.

19. Pandey A, Galvani AP. The global burden of HIV and prospects for control. The Lancet HIV 2019; 6(12):e809–e811.

20. Frank TD, Carter A, Jahagirdar D, Biehl MH, Douwes-Schultz D, Larson SL, et al. Global, regional, and national incidence, prevalence, and mortality of HIV, 1980–2017, and forecasts to 2030, for 195 countries and territories: a systematic analysis for the Global Burden of Diseases, Injuries, and Risk Factors Study 2017. The Lancet HIV 2019; 6(12):e831–e859.

